# Association between clinical characteristics and laboratory findings with outcome of hospitalized COVID-19 patients, a report from northeast of Iran

**DOI:** 10.1101/2021.01.23.21250359

**Authors:** Sahar Sobhani, Reihaneh Aryan, Elham Kalantari, Salman Soltani, Nafise Malek, Parisa Pirzadeh, Amir Yarahmadi, Atena Aghaee

**Author notes:** Correspondence should be addressed to Atena Aghaee.

## Abstract

Coronavirus disease 2019 (COVID-19) was first discovered in December 2019 in China and has rapidly spread worldwide. Clinical characteristics, laboratory findings, and their association with the outcome of patients with COVID-19 can be decisive in management and early diagnosis. Data were obtained retrospectively from medical records of 397 hospitalized COVID-19 patients between February and May 2020 in Imam Reza hospital, northeast of Iran. Clinical and laboratory features were evaluated among survivors and non-survivors. The correlation between variables and duration of hospitalization and admission to the Intensive Care Unit (ICU) was determined. Male sex, age, hospitalization duration, and admission to ICU were significantly related to mortality rate. Headache was a more common feature in patients who survived (*p* = 0.017). It was also related to a shorter stay in the hospital (*p* = 0.032) as opposed to patients who experienced chest pain (*p* = 0.033). Decreased levels of consciousness and dyspnea were statistically more frequent in non-survivors (*p* = 0.003 and *p* = 0.011, respectively). Baseline white blood cell count (WBC), erythrocyte sedimentation rate (ESR), and C-reactive protein (CRP) were significantly higher in non-survivors (*p* < 0.001). Patients with higher WBC and CRP levels were more likely to be admitted to ICU (*p* = 0.009 and *p* = 0.001, respectively). Evaluating clinical and laboratory features can help clinicians find ways for risk stratifying patients and even make predictive tools. Chest pain, decreased level of consciousness, dyspnea, and increased CRP and WBC levels seem to be the most potent predictors of severe prognosis.

## 1. Introduction

In December 2019, severe acute respiratory syndrome coronavirus (SARS-CoV-2) or coronavirus disease 2019 (COVID-19) was first discovered in Wuhan, Hubei Province, China, and has rapidly spread all over the world, causing a pandemic. The virus is considered to be transmitted by respiratory droplets and contaminated surfaces (Huang, Wang, et al. 2020, Zhu et al. 2020, Alamdari et al. 2020). As of October 2020, there are >1 million documented deaths caused by COVID-19 (JPA. October 7, 2020.). In Iran, on February 19, 2020, two patients in Qom city were confirmed as COVID-19 positive. Ultimately, the disease spread expeditiously in adjacent provinces near Qom, such as Tehran, Markazi, Isfahan, and Khorasan Razavi provinces, and shortly after, in all 31 provinces of the country (JPA. October7, 2020.). To help better diagnose COVID-19, clinicians, and public health professionals should consider the wide variety of signs, symptoms, and laboratory findings of COVID-19 (Abdi 2020).

Although COVID-19 has various clinical manifestations, most patients experience very little or no symptoms, especially in the early disease stage (Alhazzani et al., Wang et al. 2020). The incubation period of COVID-19, extending from exposure to onset of disease symptoms, is estimated at approximately 5.2 days (Bai et al. 2020, Alamdari et al. 2020). This infection’s common signs and symptoms include fever, cough, fatigue, sore throat, chest pain, dyspnea, myalgia, headache, anosmia, ageusia, and diarrhea. In more severe cases, the infection can cause pneumonia, acute respiratory distress syndrome (ARDS), kidney failure, and even death (Organization 2020, Wang, Tang, and Wei 2020).

The current knowledge surrounding clinical characteristics, laboratory findings, and their association in critically ill patients with COVID-19 infection is limited but can prove to be decisive in the management and early diagnosis of this deadly disease.

In this survey, we studied patients with confirmed COVID-19 who were admitted to Imam Reza Hospital of Mashhad, Iran. The data regarding the association between clinical presentations, laboratory findings, and survival of COVID-19 patients will be of considerable value for the early identification of individuals at risk of becoming critically ill and most likely to benefit from intensive care treatment.

## 2. Methods and Materials

### 2.1. Study population and data collection

All patients with confirmed COVID-19 diagnoses admitted to Imam Reza hospital in northeast of Iran between February and May 2020 were enrolled in the study. The COVID-19 infection was confirmed by Polymerase Chain Reaction (PCR) test and lung High-Resolution Computed Tomography (HRCT) results, as instructed in Iranian national guidelines. The patient outcome was defined as the patients’ status at discharge, whether the patient was alive or deceased. All patients were admitted following moderate to severe stages of the disease (i.e., more than 40% of the lung parenchyma affected or instability of vital signs such as hypoxemia and hypotension or severe leukopenia).

In the first hours of admission, all patients were interviewed. A complete medical history, drug history, patient’s characteristics, laboratory tests, medications prescribed to the patient, para-clinical assessments, and patient outcomes were recorded in the COVID-19 registry of Imam Reza Hospital. Patients were treated according to the national and universal COVID-19 management guidelines.

### 2.2. Ethics statement

The study protocol was approved by the Ethics Board of Mashhad University of Medical Sciences (IR.MUMS.REC.1399097). Patient’s personal information was entered as codes in the database, and the identities of patients were anonymous. The study was done under the principles of the Declaration of Helsinki.

### 2.3. Statistical Analysis

Categorical variables were expressed as frequency and percentage. Moreover, continuous variables were expressed as mean ± standard deviation (SD). Chi-squared test, Fisher exact test, and Student’s t-test were used for nonparametric and parametric analysis, respectively. Pearson’s correlation test was used to assess the relationship between clinical features and laboratory findings with hospitalization duration and admission to ICU. A *p*-value < 0.05 was considered statistically significant. All statistical analyses were performed using SPSS 22 software (SPSS, Chicago, IL, USA).

## 3. Results

A total of 397 patients were included in this analysis. The mean age was 60.6 years (ranging from 14 to 94 years), 223 (56.2%) were males, and 174 (43.8%) were females. 336 patients (84.6%) survived and were discharged in a stable condition, but unfortunately, 61 patients (15.4%) were deceased. The mean duration of hospitalization was 9.3 days, which was significantly longer in the non-survivors (*p* < 0.001). The mean age was higher in those who died than those discharged alive (66.01 and 59.64 years, respectively) (*p* = 0.013). Males had a higher mortality rate compared to females (18.8% and 10.9%, *p* = 0.035). 68 patients (17.5%) required ICU management, and mortality was significantly higher in these patients (*p* < 0.001) (Table 1).

**Table 1.**
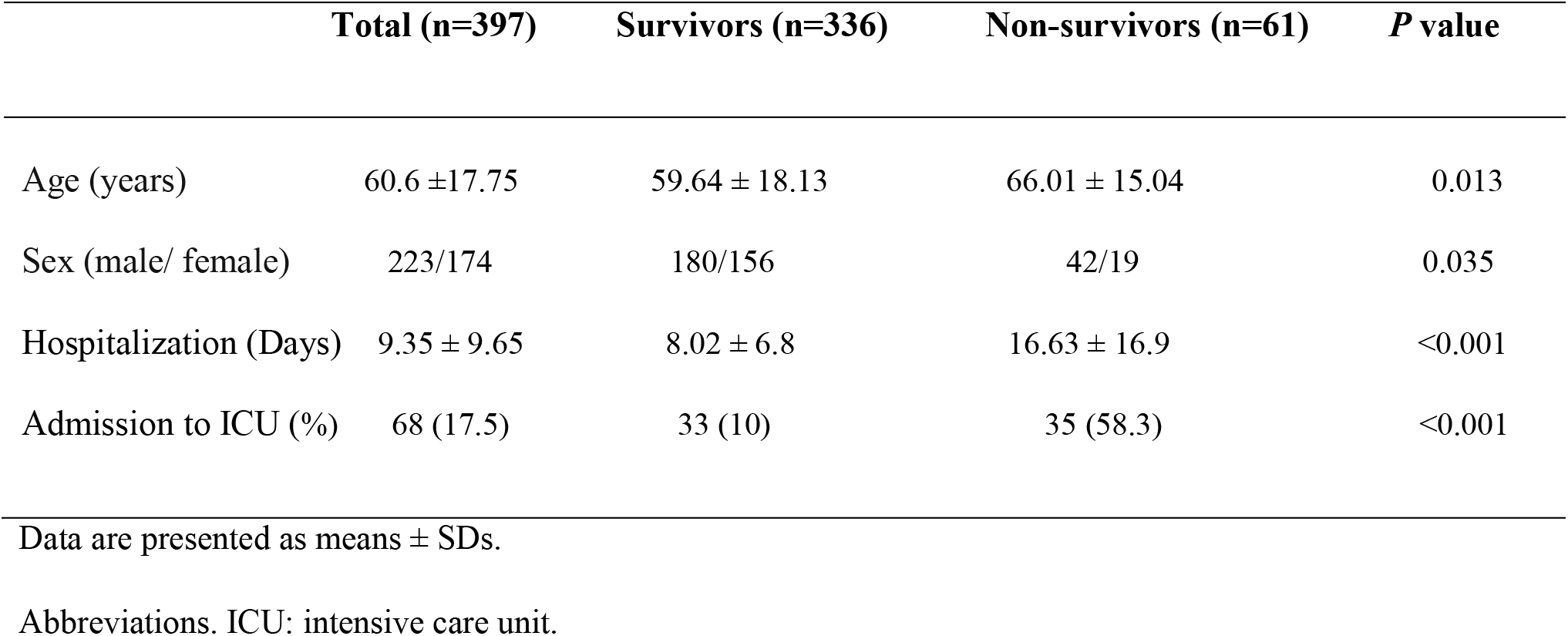
Characteristics of survivors and non-survivors COVID-19 patients.

|  | **Total (n=397)** | **Survivors (n=336)** | **Non-survivors (n=61)** | ***P* value** |
| --- | --- | --- | --- | --- |
| Age (years) | 60.6 ±17.75 | 59.64 ± 18.13 | 66.01 ± 15.04 | 0.013 |
| Sex (male/ female) | 223/174 | 180/156 | 42/19 | 0.035 |
| Hospitalization (Days) | 9.35 ± 9.65 | 8.02 ± 6.8 | 16.63 ± 16.9 | <0.001 |
| Admission to ICU (%) | 68 (17.5) | 33 (10) | 35 (58.3) | <0.001 |

Data are presented as means ± SDs.
Abbreviations. ICU: intensive care unit.

Common baseline clinical signs/symptoms among our patients were chest tightness (261 patients [65.7%]), fever (225 patients [56.7%]) and myalgia (126 patients [31.7%]). Decreased levels of consciousness and dyspnea were statistically more frequent in non-survivors (*p* = 0.003 and *p* = 0.011, respectively). Interestingly, the headache was a more common feature in patients who survived (*p* = 0.017). We also investigated three important laboratory findings in all patients; white blood cell count (WBC), erythrocyte sedimentation rate (ESR), and C-reactive protein (CRP), which all three were significantly higher in non-survivors (*p* < 0.001) (Table 2).

**Table 2.** Baseline clinical manifestations and laboratory findings of survivors and non-survivors COVID-19 patients.

|  | Total (n=397) | Survivors (n=336) | Non-survivors (n=61) | *P* value |
| --- | --- | --- | --- | --- |
| Fever (%) | 225 (56.7) | 189 (56.3) | 32 (52.4) | 0.573 |
| Headache (%) | 59 (14.9) | 56 (16.7) | 3 (5) | 0.017 |
| Sore Throat (%) | 43 (10.8) | 36 (10.7) | 7 (11.4) | 0.825 |
| Chest Pain (%) | 56 (14.1) | 51 (15.2) | 5 (8.2) | 0.166 |
| Myalgia (%) | 126 (31.7) | 110 (32.7) | 16 (26.2) | 0.369 |
| Smell Disorder (%) | 29 (7.3) | 27 (8) | 2 (3.3) | 0.284 |
| Taste Disorder (%) | 24 (6) | 22 (6.5) | 2 (3.3) | 0.557 |
| Pharyngeal Exudates (%) | 7 (1.8) | 6 (1.8) | 1 (1.6) | 0.431 |
| Decreased Consciousness (%) | 8 (2) | 3 (0.9) | 5 (8.2) | 0.003 |
| Dyspnea (%) | 74 (18.6) | 55 (16.4) | 19 (31.1) | 0.011 |
| Chest Tightness (%) | 261 (65.7) | 211 (62.8) | 43 (70.5) | 0.303 |
| Nausea and Vomiting (%) | 99 (24.9) | 87 (25.9) | 11 (18) | 0.200 |
| Diarrhea (%) | 51 (12.8) | 46 (13.7) | 5 (8.2) | 0.300 |
| Initial WBC ($10^3/\text{mm}^3$) | $8.80 \pm 5.29$ | $8.25 \pm 4.51$ | $11.67 \pm 7.67$ | <0.001 |
| Initial CRP ($\mu\text{g/dL}$) | $90.13 \pm 77.73$ | $78.86 \pm 66.80$ | $140.94 \pm 100.92$ | <0.001 |
| Initial ESR (mm/h) | $51.55 \pm 32.46$ | $50.34 \pm 32.51$ | $57.03 \pm 32.03$ | <0.001 |

Data are presented as % and means ± SDs.
Abbreviations. WBC: white blood cells; CRP: C-reactive protein; ESR: erythrocyte sedimentation rate.

We also evaluated the correlation between clinical features and laboratory findings with hospitalization duration and admission to ICU. Patients who reported headaches had a shorter stay in the hospital (*p* = 0.032) than patients who experienced chest pain, which spent more days hospitalized (*p* = 0.033). Moreover, patients with chest tightness were more likely to be admitted to ICU (*p* = 0.033) (Table 3). The patients with higher CRP levels had a longer stay in the hospital (*p* = 0.006). Furthermore, patients with higher WBC and CRP levels were more likely to be admitted to ICU (*p* = 0.009 and *p* = 0.001, respectively).

**Table 3.** Pearson’s correlation coefficients between clinical manifestations and laboratory findings with hospitalization duration and admission to ICU.

|  | Duration of Hospitalization |  | Admission to ICU |  |
| --- | --- | --- | --- | --- |
|  | *r* values | *P* values | *r* values | *P* values |
| Fever | 0.035 | 0.489 | -0.009 | 0.865 |
| Headache | -0.109 | 0.032* | 0.081 | 0.109 |
| Sore Throat | 0.013 | 0.794 | -0.033 | 0.520 |
| Chest Pain | 0.108 | 0.033* | -0.015 | 0.765 |
| Myalgia | 0.001 | 0.983 | 0.043 | 0.397 |
| Smell Disorder | 0.027 | 0.597 | -0.002 | 0.972 |
| Taste Disorder | 0.043 | 0.403 | -0.034 | 0.508 |
| Pharyngeal Exudates | 0.0005 | 0.996 | 0.040 | 0.437 |
| Decreased Consciousness | -0.008 | 0.875 | 0.089 | 0.078 |
| Dyspnea | 0.037 | 0.462 | 0.070 | 0.168 |
| Chest Tightness | 0.021 | 0.679 | 0.108 | 0.033* |
| Nausea and Vomiting | -0.029 | 0.568 | 0.018 | 0.729 |
| Diarrhea | -0.013 | 0.791 | 0.042 | 0.411 |
| Initial WBC ($10^3/\text{mm}^3$) | -0.002 | 0.975 | 0.139 | 0.009** |
| Initial CRP ($\mu\text{g/dL}$) | 0.16 | 0.006** | 0.213 | 0.001** |
| Initial ESR (mm/h) | 0.045 | 0.454 | 0.073 | 0.228 |

Data are presented as means ± SDs.
\*Correlation is significant at the 0.05 level.
\*\*Correlation is significant at the 0.01 level.
Abbreviations. WBC: white blood cells; CRP: C-reactive protein; ESR: erythrocyte sedimentation rate.

## 4. Discussion

COVID-19 has a range of severity from no symptoms to ARDS. The specific determining factors for the severity of the disease are not well recognized yet. Regarding limited health facilities and a large number of patients referred to hospitals, knowing more about this disease’s clinical course helps decide whom to be admitted. In other words, the more we know about the determinants of the clinical course, we can use them in risk stratifying patients and predicting clinical outcomes. In the present study, the mean age of patients was 60.6 years old. A report from the United States in March 2020 showed that 62% of COVID-19 patients were older than 55 (COVID TC). In our study, the mean age of non-survivors was significantly higher than those who survived (66.01 and 59.64, respectively). This finding was also noted in some other studies, such as in Italy (Cecconi et al. 2020) and United States (Imam et al. 2020). They found older age as a predictor of mortality in patients with COVID-19. Our results showed that the mortality rate was higher in males. Similarly, in a cohort study by Palaidimos L et al. in the United States, male sex was associated with worse outcomes (Palaiodimos et al. 2020). Another study in Wuhan also showed male sex as a risk factor for severity and mortality (Chen, Bai, et al. 2020).

WBC may be useful in risk stratifying. In our study, the initial WBC count was in direct correlation with mortality and ICU admission. Unfortunately, we did not include lymphocyte count in this survey. Several studies showed that lymphocyte percentage is inversely associated with the severity of COVID-19 disease (Tan, Wang, et al. 2020, Tan, Huang, et al. 2020). A study by Chen R et al. showed that an increased number of neutrophils was a predictor of disease severity (Chen, Bai, et al. 2020).

According to our findings, CRP levels were significantly higher in non-survivors. Also, hospitalization duration in patients with higher CRP levels was longer, and they were more likely to be admitted to ICU. Moreover, elevated CRP correlation with disease severity has been shown in other studies (Huang, Pranata, et al. 2020).

Belvis R et al. showed that headache was the fifth most frequent symptom after fever, cough, myalgia/fatigue, and dyspnea. In the present study, the headache was not a common symptom among our patients (14.9%), but the prevalence of headaches was significantly higher in survivors (*p* = 0.017). Furthermore, patients with headaches had a shorter duration of hospitalization than patients who experienced chest pain. Similarly, Trigo J et al. showed that headache is an independent predictor of lower mortality risk in COVID-19 patients (Trigo et al. 2020). In another study, evaluating 179 hospitalized patients with COVID-19, mortality was shown to be higher in patients with headache, although it was not statistically significant in multivariate regression analysis (Du et al. 2020).

In a study about the symptoms of COVID-19 patients in the United States, chest pain was among the symptoms reported more commonly after March 8, 2020 (8% before and 35% after). However, a more significant portion of patients in this report was not hospitalized (Burke et al. 2020). In our study, chest pain prevalence was not significantly different between survivors and non-survivors, but the patients who experienced chest pain spent more days in the hospital. Dyspnea, as a subjective experience of breathing discomfort, has been reported to affect less than 50% of COVID-19 patients and is more common in those who will die compared to those who recover (Chen, Wu, et al. 2020). Also, in the present study, dyspnea was more common in non-survivors than the survivor group (*p* = 0.011), keeping in mind that dyspnea seems to be underreported in patients with covid-19, which is called happy hypoxemia (Couzin-Frankel 2020).

A decreased level of consciousness is caused by a variety of etiologies, from hypoxia to neurologic complications. In our study, a decreased level of consciousness, regardless of its etiology, was more common in non-survivor. Impaired consciousness has been reported in 7.5% of hospitalized patients with covid-19, and it has been more likely to be presented in severely affected patients (Mao et al. 2020). Similarly, Lahiri D et al. reported that impaired consciousness was considered among the presenting features of COVID-19 (Lahiri and Ardila 2020).

## 5. Conclusions

The COVID-19 pandemic poses many challenges, and it is essential to gather as much knowledge as possible about this disease. Clinical characteristics, laboratory findings, and their association with the outcome of patients with COVID-19 can be decisive in the management and early diagnosis. Besides, considering the importance of optimized use of limited health facilities, it is essential to know how to stratify patients and even make predictive tools for better treatment. Chest pain, decreased level of consciousness, dyspnea, and increased CRP and WBC levels seem to be the most potent predictors of severe prognosis.

## Data Availability

All data used to support the findings of this study are included within the article.

## Data Availability

All data used to support the findings of this study are included within the article.

## Author Declarations

We confirm all relevant ethical guidelines have been followed, and any necessary IRB and/or ethics committee approvals have been obtained.

## Acknowledgments

This study was supported by the Vice-Chancellor for research affairs and Research Hospital Ethics Committee of Mashhad University of Medical Sciences (Grant number: 990081).

